# Comparison of first trimester dating methods for gestational age estimation and their implication on preterm birth classification in a North Indian cohort

**DOI:** 10.1101/2019.12.27.19016006

**Authors:** Ramya Vijayram, Nikhita Damaraju, Ashley Xavier, Bapu Koundinya Desiraju, Ramachandran Thiruvengadam, Sumit Misra, Shilpa Chopra, Ashok Khurana, Nitya Wadhwa, GARBH-Ini Study Group, Raghunathan Rengaswamy, Himanshu Sinha, Shinjini Bhatnagar

**Author notes:** Correspondence: Shinjini Bhatnagar, Maternal and Child Health Program, Translational Health Science and Technology Institute, Faridabad, India; Himanshu Sinha, Department of Biotechnology, Bhupat and Jyoti Mehta School of Biosciences, Indian Institute of Technology Madras, Chennai, India. Ramya Vijayram, Nikhita Damaraju, and Ashley Xavier are joint first authors.

## Abstract

**Background:** Different formulae have been developed globally to estimate gestational age (GA) by ultrasonography in the first trimester of pregnancy. In this study, we develop an Indian population-specific dating formula and compare its performance with published formulae. Finally, we evaluate the implications of the choice of dating method on preterm birth (PTB) rate. This study’s data was from GARBH-Ini, an ongoing pregnancy cohort of North Indian women to study PTB.

**Methods:** Comparisons between ultrasonography-Hadlock and last menstrual period (LMP) based dating methods were made by studying the distribution of their differences by Bland-Altman analysis. Using data-driven approaches, we removed data outliers more efficiently than by applying clinical parameters. We applied advanced machine learning algorithms to identify relevant features for GA estimation and developed an Indian population-specific formula (Garbhini-GA1) for the first trimester. PTB rates of Garbhini-GA1 and other formulae were compared by estimating sensitivity and accuracy.

**Results:** Performance of Garbhini-GA1 formula, a non-linear function of crown-rump length (CRL), was equivalent to published formulae for estimation of first trimester GA (LoA, - 0.46,0.96 weeks). We found that CRL was the most crucial parameter in estimating GA and no other clinical or socioeconomic covariates contributed to GA estimation. The estimated PTB rate across all the formulae including LMP ranged 11.27 – 16.50% with Garbhini-GA1 estimating the least rate with highest sensitivity and accuracy. While the LMP-based method overestimated GA by three days compared to USG-Hadlock formula; at an individual level, these methods had less than 50% agreement in the classification of PTB.

**Conclusions:** An accurate estimation of GA is crucial for the management of PTB. Garbhini-GA1, the first such formula developed in an Indian setting, estimates PTB rates with higher accuracy, especially when compared to commonly used Hadlock formula. Our results reinforce the need to develop population-specific gestational age formulae.

## Background

Preterm birth (PTB) is conventionally defined as a birth that occurs before 37 completed weeks of gestation [1,2]. Globally, complications arising from preterm birth were the leading cause of child (less than 5 years of age) mortality in 2016, accounting for 35% of neonatal deaths [3]. PTB is a unique disease in the way it is defined by the duration of gestation and not by a pathological process. The duration of gestation is the period between the date of conception and date of delivery. While the date of delivery can be documented with fair accuracy, ascertaining the date of conception is challenging. The estimation of gestational age (GA) during the antenatal period also called as the dating of pregnancy has been conventionally done using the first day of the recall-based last menstrual period (LMP) or measurement of foetal biometry by ultrasonography (USG) [4,5]. Each of these methods poses a unique set of challenges. The accuracy of dating by LMP method is dependent on accurate recall, and regularity of menstrual cycle [4,6] which, is affected by numerous physiological and pathological conditions such as obesity [7], polycystic ovarian syndrome [8], breastfeeding [9] and use of contraceptive methods [10].

The USG method is based on foetal biometry using crown-rump length (CRL) in the first trimester. Several formulae exist to estimate GA using CRL, including Hadlock formula [11], based on a US population-based study widely used in India [12]. However, the choice of dating formula might influence dating accuracy, as these formulae have been developed from studies that differed both in the study population and study design [13]. The error and bias due to the choice of dating formula need to be quantitatively studied to estimate the rate of PTB in a specific population [14]. In addition to its public health importance, accurate dating is essential for clinical decision making during the antenatal period, such as scheduling monitoring visits and recommending appropriate antenatal care [4].

This study first quantified the discrepancy between LMP and USG-based (Hadlock) dating methods during the first trimester in an Indian population. We characterised how each method could contribute to the discrepancy in calculating the GA. We then built a population-specific model from the GARBH-Ini cohort (Interdisciplinary Group for Advanced Research on BirtH outcomes - DBT India Initiative), Garbhini-GA1, and compared its performance with the published ‘high quality’ formulae for the first-trimester dating [13] – McLennan and Schluter [15], Robinson and Fleming [16], Sahota [17] and Verburg [18], INTERGROWTH-21^st^ [19], and Hadlock’s formula [11] (Table S1). Finally, we quantified the implications of the choice of dating methods on PTB rates in our study population.

## Methods

### Study design

GARBH-Ini is a collaborative program, initiated by Translational Health Science and Technology Institute, Faridabad with partners from Regional Centre of Biotechnology, Faridabad; National Institute of Biomedical Genomics, Kalyani; Civil Hospital, Gurugram; Safdarjung hospital, New Delhi. The GARBH-Ini cohort is a prospective observational cohort of pregnant women initiated in May 2015 at the District Civil Hospital that serves a mostly rural and semi-urban population in the Gurugram district, Haryana, India. The cohort study’s objective is to develop an effective risk stratification that facilitates timely referral for women at high risk of PTB, particularly in low- and middle-income countries. Women in the GARBH-Ini cohort are enrolled within 20 weeks of gestation and are followed three times during pregnancy till delivery and once postpartum [20]. After a verbal consent to be interviewed, informed consent for screening is obtained for women at < 20-weeks of gestational age (GA) calculated by the last menstrual period. A dating ultrasound is performed within the week to confirm a viable intrauterine pregnancy with < 20-weeks GA using standard foetal biometric parameters. A time-series data on a large set of clinical and socioeconomic variables are collected across pregnancy to help stratify women into defined risk groups for PTB. The dating ultrasound is performed by a qualified radiologist specifically trained in the study protocol. The clinical and demographic information is collected by trained, dedicated research staff under medically qualified research officers’ supervision. The data acquisition protocols and quality control measures are detailed elsewhere [20].

### Sampling strategy and participant datasets derived for the study

This analysis’s samples were derived from the first 3499 participants enrolled in the GARBH-Ini study (between May 2015 to November 2017). We included 1721 participants (N_p_ = 1721), enrolled < 14 weeks of gestation and who had information on the LMP, CRL with singleton pregnancy which advanced beyond 20 weeks of gestation, i.e. the pregnancy did not end in a spontaneous abortion or major congenital abnormalities which required medical termination of pregnancy. If a participant was enrolled < 11 weeks, dating ultrasound was done upon enrolment when CRL was measured for the first time. The same participant was asked to come for another ultrasound between 11-14 weeks of gestation to assess foetal morphology during which another CRL measurement was taken. If more than one scan was performed for a participant, data from both the scans were included as unique observations (N_o_). Therefore, 1721 participants contributed a total of 2562 observations (N_o_ = 2562) that was used for further analyses, and this dataset of observations was termed as the TRAINING DATASET (Figure 1). This dataset was used to develop a population-based dating model named Garbhini-GA1, for the first trimester.

**Figure 1:**
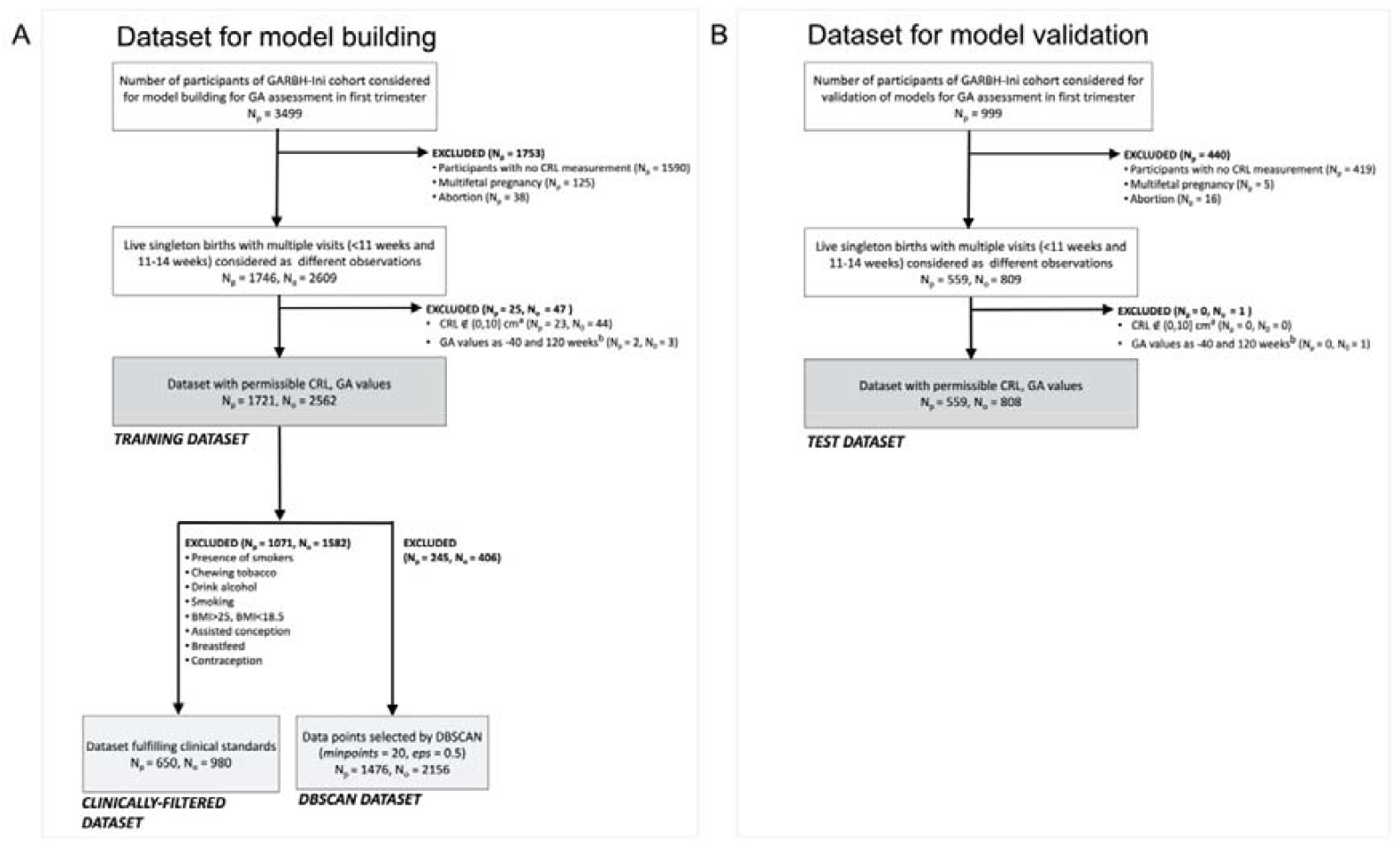
Outline of the data selection process for different datasets – (A) TRAINING DATASET and (B) TEST DATASET. Coloured boxes indicate the datasets used in the analysis. The names of each of the dataset are indicated below the box. Exclusion criteria for each step are indicated. N_p_ indicates the number of participants included or excluded by that particular criterion and N_o_ indicates the number of unique observations derived from the participants in a dataset. Biologically implausible CRL values (either less than 0 or more than 10 cm) for the first trimester were excluded, b Biologically implausible GA values (either less than 0 and more than 45 weeks) were excluded.

It is essential to independently evaluate models on data that was not used for building the model in order to eliminate any biases that may have been incorporated due to the iterative learning process of the model building dataset and estimate the expected performance when applying the model on new data in the real world. We used an unseen TEST DATASET created from 999 participants enrolled after the initial set of 3499 participants in this cohort (Figure 1). The TEST DATASET was obtained by applying identical processing steps as described for the TRAINING DATASET (N_o_ = 808 from N_p_ = 559; Figure 1).

### Assessment of LMP and CRL

The date of LMP was ascertained from the participant’s recall of the first day of the last menstrual period. CRL from an ultrasound image (GE Voluson E8 Expert, General Electric Healthcare, Chicago, USA) was captured in the midline sagittal section of the whole foetus by placing the callipers on the outer margin skin borders of the foetal crown and rump ([20], see Supplementary Figure S5). The CRL measurement was done thrice on three different ultrasound images, and the average of the three measurements was considered for estimation of CRL-based GA. Under the supervision of medically qualified researchers, study nurses documented the clinical and sociodemographic characteristics [20].

### Development and validation of the population-specific gestational dating model

The gold standard or ground truth for development of first-trimester dating model was derived from a subset of participants with the most reliable GA based on last menstrual period. We used two approaches to create subsets from the TRAINING DATASET for developing the first-trimester population-based dating formula. The first approach excluded participants with potentially unreliable LMP or high risk of foetal growth restriction, giving us the CLINICALLY-FILTERED DATASET (N_o_ = 980 from N_p_ = 650; Figure 1, Table S2).

The second approach used Density-Based Spatial Clustering of Applications with Noise (DBSCAN) method to remove outliers based on noise in the data points. DBSCAN identifies noise by classifying points into clusters if there are a sufficient number of neighbours that lie within a specified Euclidean distance or if the point is adjacent to another data point meeting the criteria [21]. DBSCAN was used to identify and remove outliers in the TRAINING DATASET using the parameters for distance cut-off (*epsilon, eps*) 0.5 and the minimum number of neighbours (*minpoints*) 20. A range of values for eps and minpoints did not markedly change the clustering result (Table S3). The resulting dataset that retained reliable data points for the analysis was termed as the DBSCAN DATASET (N_o_ = 2156 from N_p_ = 1476; Figure 1).

The use of CRL for dating of pregnancy is restricted to the first trimester of pregnancy in clinical practice. This is because of the technical difficulties in obtaining accurate CRL measurements beyond this period. The same was practised in the GARBH-Ini cohort as it is an observational study. When an ultrasonographic examination was performed during early pregnancy, the radiologist refrained from measuring CRL if she/he was not assured of its accuracy. Instead, the radiologist measured the other foetal biometry (biparietal diameters, abdominal and head circumference and femur length to ascertain the gestational age). This resulted in a dataset with GA by CRL truncated at 14 weeks of gestation. When used for training models, such a truncated dataset may lead to inaccuracies in the model fitting particularly at the margins of the distribution around 14 weeks [22]. We considered multiple approaches used in the literature [22] and overcame this by supplementing our dataset with simulated observations from the Hadlock dataset, which measured the relationship between CRL and GA in the range of 15 – 18 weeks [11]. This supplemented dataset was used to build fractional polynomial models of GA as a function of CRL (see Figure S1, Table S7).

Development of a first trimester dating formula was done by fitting fractional polynomial regression models of GA (weeks) as a function of CRL (cm) on CLINICALLY-FILTERED and DBSCAN datasets. The performance of the chosen formula was validated in the TEST DATASET.

In addition to CRL as a primary indicator, a list of 282 candidate variables was explored by feature selection methods on the DBSCAN DATASET to identify other variables which may be predictive of GA during the first trimester. These methods helped to find uncorrelated, non-redundant features that might improve GA prediction accuracy (Table S4). First, the feature selection was done using Boruta [23], a random forest classifier, which identified six features and second, by implementing Generalised Linear Modelling (GLM) that identified two candidate predictors of GA. A union of these features (Table S5), gave a list of six candidate predictors. Equations were generated using all combinations of these predictors in the form of linear, logarithmic, polynomial and fractional power equations. The best fit model was termed Garbhini-GA1 formula and was validated for its performance in the TEST DATASET.

### Comparison of LMP- and USG-based dating methods during the first-trimester

We calculated the difference between LMP- and USG-based GA for each participant and studied the distribution of the differences by Bland-Altman (BA) analysis [24]. Additionally, we estimated the effect of factors that could contribute to the discrepancy between GA by LMP and ultrasound. This may be due to an unreliable LMP or foetal growth restriction. We repeated the comparative analysis in our population’s subsets with accurate LMP and no risk factors for foetal growth restriction (see Additional File 1).

The mean difference between the methods and the limits of agreement (LoA) for 95% CI were reported. The PTB rates with LMP- and USG-based methods were reported per 100 live births with 95% CI. We compared different USG-based formulae using correlation analysis.

The data analyses were carried out in R versions 3.6.1 and 3.5.0. DBSCAN was implemented using the package *dbscan*, and the random forests feature selection was performed using the *Boruta* package [23]. Statistical analysis for comparing PTB rate as estimated using different dating formulae was carried out using standard t-test with or without Bonferroni multiple testing correction or using Fisher’s Exact test wherever appropriate.

## Results

### Description of participants included in the study

The median age of the participants enrolled in the cohort was 23.0 years (IQR 21.0 – 26.0), with the median weight and height as 47.0 kg (IQR 42.5 – 53.3) and 153.0 cm (IQR 149.2 – 156.8), respectively and with 59.93% of the participants having a normal first trimester BMI (median 20.09, IQR 18.27 – 22.59). Almost half of them were primigravida. Most of the participants (98.20%) were from middle or lower socioeconomic strata [25]. The participants selected for this analysis had a median GA of 11.71 weeks (IQR 9.29 – 13.0). The other baseline characteristics are given in Table 1.

**Table 1:** Baseline characteristics of the participants included in the TRAINING DATASET (N_o_ = 2562) to compare different methods of dating.

| Sociodemographic characteristics | Median (IQR) or N (%) or Mean ± SD |
| --- | --- |
| Age (year) | 23 (21 – 26) |
| GA at enrolment by LMP (weeks) | 11.31 ± 2.67 |
| GA at enrolment by USG-Hadlock (weeks) | 10.87 ± 2.28 |
| BMI at enrolment into the cohort <sup>a</sup> |  |
| Underweight | 27.20% |
| Normal weight | 59.93% |
| Obese | 9.09% |
| Overweight | 1.66% |
| Haemoglobin (g/dL) | 8.8 (8.2 – 9.2) |
| Height (cm) | 153.0 (149.2 – 156.8) |
| Socioeconomic status <sup>b</sup> |  |
| Upper class | 0.66% |
| Upper middle class | 15.40% |
| Lower middle class | 33.98% |
| Upper lower class | 48.96% |
| Lower class | 0.43% |
| Undetermined | 0.57% |
| Parity (number) |  |
| 0 | 49.53% |
| 1 | 33.55% |
| 2 | 12.60% |
| 3 | 3.34% |
| 4 | 0.74% |
| 5 | 0.14% |
| Level of education |  |
| Illiterate | 21.58% |
| Literate or primary school | 8.63% |
| Middle school | 15.09% |
| High school | 18.61% |
| Post high school diploma | 20.89% |
| Graduate | 12.23% |
| Post-graduate | 2.94% |
| Occupation |  |
| Unemployed | 93.48% |
| Unskilled worker | 3.34% |
| Semi-skilled worker | 0.97% |
| Skilled worker | 1.40% |
| Clerk, shop, farm owner | 0.17% |
| Semi-professional | 0.26% |
| Professional | 0.34% |
| Religion |  |
| Hindu | 92.14% |
| Muslim | 6.60% |
| Sikh | 0.40% |
| Christian | 0.74% |
| Buddhist | 0.00% |
| More than one religion | 0.09% |
| Fuel used for cooking <sup>c</sup> |  |
| Biomass fuel | 7.86% |
| Clean fuel <sup>d</sup> | 92.14% |
| Source of drinking water |  |
| Safe water <sup>e</sup> | 49.80% |
| Unsafe water | 50.20% |
| Second-hand tobacco smoke |  |
| Exposed | 19.23% |
| Unexposed | 80.57% |
| Undetermined | 0.20% |
| History of any chronic illnesses<sup>f</sup> |  |
| Absent | 99.03% |
| Present | 0.97% |
| History of hypertensive disease of pregnancy |  |
| Absent | 99.57% |
| Present | 0.43% |
| History of contraceptive at the time of conception |  |
| Absent | 90.79% |
| Present | 7.30% |

<sup>a</sup> Pre-pregnancy BMI was calculated as weight (kg)/height<sup>2</sup> (m) from participants' weight and height measured at enrolment. BMI categories were defined as underweight (< 18.5); normal (18.5 – 24.9); overweight (25.0 – 29.9); obese (≥ 30.0).
<sup>b</sup> Socioeconomic status was assessed using Modified Kuppuswamy's socioeconomic scale [25], calculated using education and occupation of the head of the family and monthly family income.
<sup>c</sup> Indoor air pollution: use of biomass fuel for cooking or presence of a smoker in the residential compound, as reported by the participant.
<sup>d</sup> Clean fuel includes liquefied petroleum gas and electricity.
<sup>e</sup> Safe water includes bottled water or piped water into the residence.
<sup>f</sup> Chronic illnesses include a history of hypertension, diabetes, cardiac disease and thyroid disorders.

### Comparison of USG-Hadlock and LMP-based methods for estimation of GA in the first trimester

The mean difference between USG-Hadlock and LMP-based dating at the time of enrolment was found to be −0.44 ± 2.02 weeks (Figure 2a) indicating that the LMP-based method overestimated GA by nearly three days. The LoA determined by BA analysis was −4.39, 3.51 weeks, with 8.82% of participants falling beyond these limits (Figure 2b) suggesting a high imprecision in both the methods. The LoA between USG-Hadlock and LMP-based dating marginally narrowed when tested on participants with reliable LMP (LoA −4.22, 3.28) or those with low-risk of foetal growth restriction (LoA −4.13, 3.21). The wide LoA that persisted despite ensuring reliable LMP and standardised CRL measurements represent the residual imprecision due to unknown factors in GA’s estimation.

**Figure 2:**
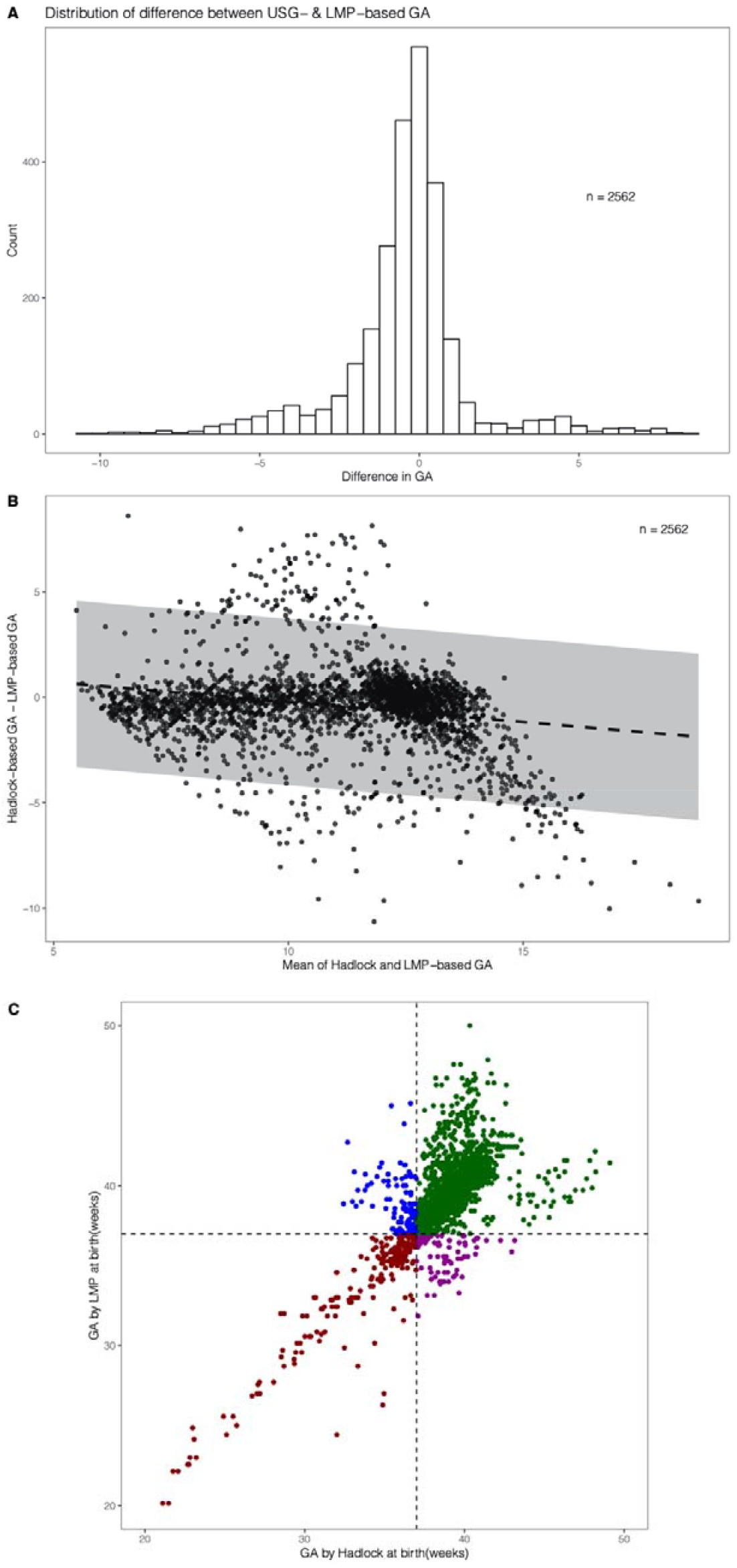
(A) Distribution of the difference between USG- and LMP-based GA. The x-axis is the difference between USG and LMP-based GA in weeks, and the y-axis is the number of observations. (B) BA analysis to evaluate the bias between USG and LMP-based GA. The x-axis is mean of Hadlock and LMP-based GA in weeks, and the y-axis is the difference between Hadlock and LMP-based GA in weeks. Regression line with 95% CI is shown. (C) Comparison of individual-level classification of preterm birth by Hadlock- and LMP-based methods. Green (term birth for both), red (preterm birth for both), blue (term birth for LMP but preterm birth for Hadlock) and purple (term for Hadlock but preterm for LMP).

### Development of Garbhini-GA1 formula for first-trimester dating

To remove noise from the TRAINING DATASET for building population-specific first-trimester dating models, two methods were used – clinical criteria-based filtering and DBSCAN (Figure 1). When clinical criteria (Figure 1) were used, more than two-third observations (68.46%) were excluded (Figure 3a). However, when DBSCAN was implemented, less than one-sixth observations (15.85%) were removed (Figure 3b). Models for first-trimester dating using CLINICALLY-FILTERED and DBSCAN datasets with CRL as the only predictor was done using fractional polynomial regression to identify the best predictive model (Figure S2). The DBSCAN approach provided a more accurate dataset (i.e. no artefacts as observed in the CLINICALLY-FILTERED DATASET) with lesser outliers. We, therefore, used DBSCAN DATASET for building dating models. Comparison among various dating models showed that the best regression coefficient (*R*^*2*^) was for quadratic regression (*R*^*2*^ = 0.86, Table S6). This provided the basis for using the following quadratic formula as the final model for estimating GA in the first trimester and was termed as Garbhini-GA1 formula:

**Figure 3:**
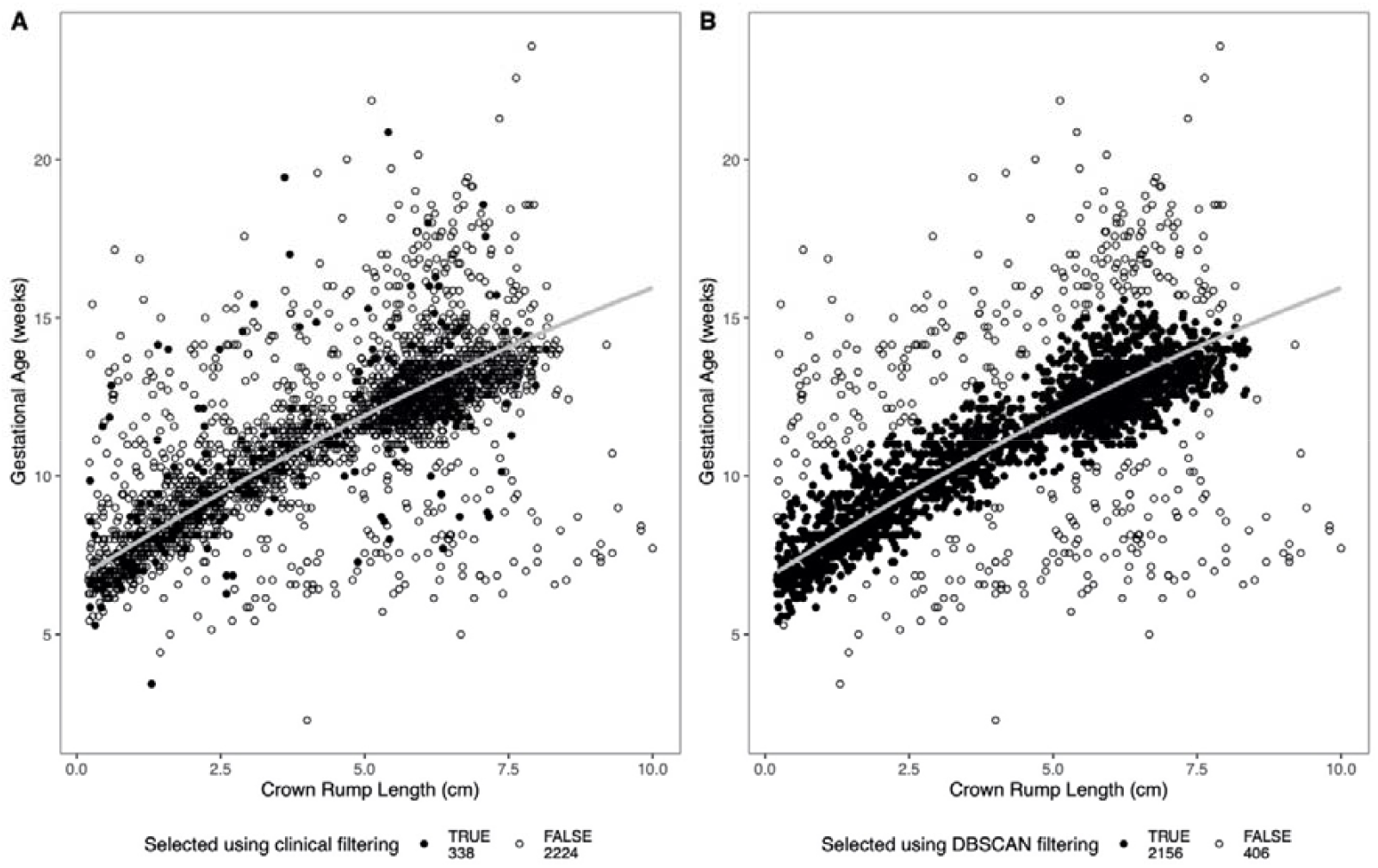
Comparison of data chosen to be reference data for the development of dating formula by (A) clinical and (B) data-driven (DBSCAN) approaches. The x-axis is CRL in cm, and the y-axis is GA in weeks (LMP-based are datapoints, Garbhini-GA1 is regression line). After filtering, the data points selected (TRUE) are coloured black and points not selected (FALSE) are white.

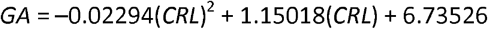

where GA is in weeks, and CRL is in cm.

A multivariate dating model including CRL and the six additional predictors identified by data-driven approaches (GLM and Random forests): resident state, weight, BMI, abdominal girth, age, and maternal education, did not improve the performance of the CRL-based dating model (Figure S3, Table S6).

### Comparison of published formulae and Garbhini-GA1 formula for estimation of GA

The actual test of the validity of a formula is to estimate GA reliably in an unseen sample population. We tested the published formulae’s performance (Table S1) and Garbhini-GA1 formula independently on the TEST DATASET (Figure S4). It was observed that Garbhini-GA1 had an *R*^*2*^ value of 0.58 (Table S8). All other formulae performed identically to Garbhini-GA1 on the TEST DATASET (Table S8). Furthermore, all possible pairwise BA analysis of these formulae (including Garbhini-GA1) showed that the mean difference of estimated GA varied from −0.17 to 0.50 weeks (Table 2). This result shows that Garbhini-GA1 performs equally well as other formulae.

**Table 2:** Pairwise comparison of mean difference (LoA) between different first-trimester dating formulae (Difference: Column formula - Row mula). Values shown in white are for the TRAINING DATASET (N_o_ = 2562) and values shown in grey are for the TEST DATASET (N_o_ = 808) (see ethods for details).

| Formula | Hadlock | McLennan-Schluter | Robinson-Fleming | Sahota | Verburg | INTERGROWTH-21<sup>st</sup> | Garbhini-GA1 |
| --- | --- | --- | --- | --- | --- | --- | --- |
| Hadlock |  | -0.16<br>(-0.40, 0.079) | -0.17<br>(-0.36, 0.016) | 0.034<br>(-0.22, 0.29) | 0.037<br>(-0.41, 0.48) | 0.079<br>(-0.54, 0.70) | 0.38<br>(-0.11, 0.87) |
| McLennan-Schluter | 0.14<br>(-0.032, 0.31) |  | -0.015<br>(-0.16, 0.13) | 0.19<br>(0.05, 0.34) | 0.20<br>(-0.10, 0.50) | 0.24<br>(-0.36, 0.83) | 0.54<br>(-0.02, 1.10) |
| Robinson-Fleming | 0.17<br>(-0.019, 0.35) | 0.024<br>(-0.095, 0.14) |  | 0.21<br>(0.082, 0.33) | 0.21<br>(-0.097, 0.52) | 0.25<br>(-0.35, 0.85) | 0.56<br>(0.02, 1.09) |
| Sahota | -0.052<br>(-0.30, 0.19) | -0.19<br>(-0.33, -0.057) | -0.22<br>(-0.35, -0.088) |  | 0.002<br>(-0.20, 0.20) | 0.044<br>(-0.46, 0.55) | 0.35<br>(-0.15, 0.85) |
| Verburg | -0.065<br>(-0.51, 0.39) | -0.21<br>(-0.52, 0.11) | -0.23<br>(-0.54, 0.08) | -0.013<br>(-0.22, 0.19) |  | 0.042<br>(-0.45, 0.53) | 0.35<br>(-0.26, 0.95) |
| INTERGROWTH<br>-21<sup>st</sup> | -0.12<br>(-0.79, 0.55) | -0.26<br>(-0.90, 0.38) | -0.28<br>(-0.94, 0.38) | -0.066<br>(-0.62, 0.49) | -0.053<br>(-0.59, 0.49) |  | 0.30<br>(-0.03, 0.64) |
| Garbhini-GA1 | -0.40<br>(-0.93, 0.13) | -0.54<br>(-1.12, 0.04) | -0.57<br>(-1.14, 0.01) | -0.35<br>(-0.87, 0.18) | -0.34<br>(-0.96, 0.29) | -0.28<br>(-0.61, 0.05) |  |

### Impact of the choice of USG dating formula on the estimation of the rate of PTB

The PTB rates estimated using different methods ranged between 11.27 and 16.5% with Garbhini-GA1 estimating the least (11.27%; CI 9.70, 13.00), followed by LMP (13.99%; CI 12.25, 15.86), Hadlock (14.53%; CI 12.77, 16.43), and Robinson-Fleming formula being the highest (16.50%; CI 14.64, 18.49). Among all pairwise comparisons performed, the differences in PTB rates estimated by Garbhini-GA1 compared with Robinson-Fleming or McLennan-Schluter were statistically significant (Fisher’s Exact test with Bonferroni correction for p < 0.05, Table S9). Furthermore, Garbhini-GA1 formula had the highest sensitivity and balanced accuracy (Table S10).

When these methods were used to determine PTB at an individual level, the Jaccard similarity coefficient (a statistic used for gauging the similarity and diversity of sample sets) ranged between 0.49 – 0.98 (Table 3). Interestingly, even though the two most used methods of dating, LMP and USG-Hadlock had similar PTB rates (13.99 and 14.53%, respectively) at the population-level, the Jaccard similarity coefficient was only 0.49 suggesting a poor agreement between the methods at an individual-level (Figure 2C, Table 3).

**Table 3:** The Jaccard similarity coefficient of PTB classification between each pair of the method.

| Formula | LMP | Hadlock | McLennan-Schluter | Robinson-Fleming | Sahota | Verburg | INTERGROWTH-21<sup>st</sup> | Garbhini-GA1 |
| --- | --- | --- | --- | --- | --- | --- | --- | --- |
| LMP | 1.00 | 0.49 | 0.50 | 0.50 | 0.52 | 0.53 | 0.53 | 0.50 |
| Hadlock |  | 1.00 | 0.90 | 0.88 | 0.88 | 0.81 | 0.80 | 0.77 |
| McLennan-Schluter |  |  | 1.00 | 0.98 | 0.83 | 0.82 | 0.80 | 0.69 |
| Robinson-Fleming |  |  |  | 1.00 | 0.82 | 0.81 | 0.79 | 0.68 |
| Sahota |  |  |  |  | 1.00 | 0.92 | 0.89 | 0.83 |
| Verburg |  |  |  |  |  | 1.00 | 0.87 | 0.83 |
| INTERGROWTH-21<sup>st</sup> |  |  |  |  |  |  | 1.00 | 0.87 |
| Garbhini-GA1 |  |  |  |  |  |  |  | 1.00 |

## Discussion

### Principal findings

This study’s primary objectives were to compare different methods and formulae used for GA estimation during the first trimester, develop a population-specific dating model for the first trimester, and study the differences in PTB rate estimation using these formulae. Our findings show that the LMP-based method overestimates GA by three days compared to the USG (Hadlock) method. While this bias does not impact at the population level with similar overall PTB rates determined by both methods, interestingly, there is less than 50% agreement between these methods on who are classified as preterm at an individual level.

This is consistent with the pattern observed in a recent study from a Zambian cohort [26]. The Hadlock formula for USG-based estimation of GA was developed on a Caucasian population and has been used for several decades globally [12]. We developed and tested population-specific dating formula to estimate GA in an Indian setting. The CRL-based Garbhini-GA1 formula performed the best and addition of other clinical and sociodemographic predictors identified from machine learning tools did not improve the performance of CRL-based Garbhini-GA1 formula. While most of the dating formulae estimated similar PTB rates, Garbhini-GA1 formula estimated the lowest PTB rate and had the best sensitivity to determine preterm birth.

### Strengths of the study

The Garbhini-GA1 formula developed from Indian population overcomes the low representativeness of existing dating formulae. Using advanced data-driven approaches, we evaluated multiple combinations of various clinical and sociodemographic parameters to estimate gestational age. We conclusively show that CRL is the sufficient parameter for first-trimester dating of pregnancy and the addition of other clinical or social parameters do not improve the performance of the dating model. Further, to build Garbhini-GA1 formula, we used a data-driven approach to remove outliers that retained more observations for building the model than would have been possible if the clinical criteria-based method had been used to develop the reference standard. Another important strength of our study is the standardised measurement of CRL. This reduces the imprecision to the minimum and makes USG-based estimation of gestational age accurate.

### Limitations of the data

For the development of Garbhini-GA1 model, it would have been ideal to have used documented LMP collected pre-conceptionally. Since our GARBH-Ini cohort enrols participants in the first trimester of pregnancy, clinical criteria based on data collected using a questionnaire was used to derive a subset of participants with reliable LMP. This was relatively incomplete as we had residual imprecision, which was not accounted for by the clinical criteria. We tried to overcome this limitation by using data-driven approaches to improve precision.

To address the truncation problem [22], we supplemented observations simulated from Hadlock distribution. While it is possible that the supplemented data points from the Hadlock formula could be different from our population data, since CRL is not measured beyond 14 weeks as standard clinical practice, this is the best possible way to address this issue.

### Interpretation

The LMP-based dating is prone to errors from recall and irregularity of menstrual cycles due to physiological causes and pathological conditions. The overestimation of GA by the LMP-based method seen in our cohort has been reported in other populations from Africa and North America [26,27]. However, the magnitude of overestimation varies, as seen in studies done earlier [26-28]. These differences could be attributed to the precision and accuracy with which these cohorts’ participants recalled their LMP. In our study, the bias in LMP-based dating was not reflected in the population-level PTB rates; however, at an individual level, LMP and USG-Hadlock had less than 50% agreement in the classification of PTB. Such considerable discordance is concerning as the clinical decisions during the early neonatal period largely depend on GA at birth. Further, any clinical and epidemiological research studying the risk factors and complications of PTB will be influenced by choice of dating method.

As shown by BA analysis, Garbhini-GA1 formula based on first-trimester CRL of our study population can be interchangeably used with Hadlock, INTERGROWTH-21^st^, Verburg and Sahota but not with McLennan-Schluter and Robinson-Fleming formulae. We get similar GA estimates using Hadlock, INTERGROWTH-21st, Verburg and Sahota formulae, which indicates that GA estimate using CRL is robust. However, even minimal difference in GA estimation leads to significantly different preterm estimates. The higher sensitivity of Garbhini-GA1 formula to classify PTB in our study population is encouraging but should be externally validated in other populations within the country before it can be recommended for application. It would be useful to evaluate the performance of population-specific formulae for second and third trimesters of gestation as ethnic differences in foetal growth might manifest more during this period.

## Conclusions

LMP overestimates GA by three days compared to USG-Hadlock method, and only half of the preterm birth were classified correctly by both these methods. CRL-based USG method is the best for GA estimation in the first trimester, and the addition of clinical and demographic features does not improve its accuracy. Garbhini-GA1 formula is an Indian-population based formula for estimating GA in the first trimester based on CRL as the prime parameter. It has better sensitivity than the more commonly used Hadlock formula in estimating the PTB rate. Our results reinforce the need to develop population-specific GA formulae. These results need to be further validated in subsequent multi-ethnic cohorts before being applied for broader use.

## Data Availability

All supplementary data used in the manuscript is submitted. Primary data can be shared according to the GARBH-Ini data sharing policy which available on request.

## List of Abbreviations

LMP: Last Mensural Period
GA: Gestational Age
CRL: Crown Rump Length
PTB: Preterm Birth Rate
USG: Ultrasonography
CI: Confidence Interval
GLM: Generalised Linear Model
LoA: Limits of Agreement
BA: Bland-Altman
IQR: Inter-Quartile Range
BMI: Body Mass Index

## Declarations

## Ethics approval and consent to participate

Ethics approvals were obtained from the Institutional Ethics Committees of Translational Health Science and Technology Institute; District Civil Hospital, Gurugram; Safdarjung Hospital, New Delhi (ETHICS/GHG/2014/1.43); and Indian Institute of Technology Madras (IEC/2019-03/HS/01/07). Written informed consent was obtained from all study participants enrolled in the GARBH-Ini cohort. For an illiterate woman, details of the study were explained in the presence of a literate family member or a neighbour who acted as the witness; a verbal consent and a thumb impression were taken from her along with the signature of the witness. All the methods were performed in accordance with the relevant guidelines and regulations.

## Consent for publication

Not applicable

## Availability of data and materials

The datasets used or analysed during the current study are available from the corresponding author on reasonable request. All the codes used for this paper are available at https://github.com/HimanshuLab/GARBH-Ini_GA1

## Authors’ contributions

R.T., H.S. and S.B. conceived this study, R.V., N.D. and A.X., performed data and statistical analyses, K.D. and R.T. performed data exports and contributed to data analysis, A.K., N.W., S.M. S.C. and GARBH-Ini Study Group developed and implemented the clinical data collection methods and data management in GARBH-Ini cohort, R.R. provided critical feedback on data analysis, K.D., R.T., H.S. and S.B. interpreted the results, R.V., N.D., A.X., R.T. and H.S. wrote the first draft of the manuscript and all listed authors critically revised and edited subsequent manuscript drafts. All authors approved the final draft of the manuscript.

## Acknowledgements

We thank all the participants of GARBH-Ini study. We thank Karthik Raman and Nirav Bhatt from Department of Biotechnology, Bhupat and Jyoti Mehta School of Biosciences and Initiative of Biological Systems Engineering, IIT Madras, Gagandeep Kang from Christian Medical College, Vellore, and Ashok Venkitaraman, National University of Singapore, Singapore, for their valuable suggestions.

## Members of GARBH-Ini Study Group

Translational Health Science and Technology Institute, NCR Biotech Cluster, Faridabad, India-Coordinating Institute (Vineeta Bal, Shinjini Bhatnagar (PI), Bhabatosh Das, Mahadev Dash, Bapu Koundinya Desiraju, Pallavi Kshetrapal, Sumit Misra, Uma Chandra Mouli Natchu, Satyajit Rath, Kanika Sachdeva, Dharmendra Sharma, Amanpreet Singh, Shailaja Sopory, Ramachandran Thiruvengadam, Nitya Wadhwa); National Institute of Biomedical Genomics, Kalyani, West Bengal, India (Arindam Maitra, Partha P Majumder (Co-PI) Souvik Mukherjee); Regional Centre for Biotechnology, NCR Biotech Cluster, Faridabad, India (Tushar K Maiti); Clinical Development Services Agency, Translational Health Science and Technology Institute, NCR-Biotech Cluster, Faridabad, India (Monika Bahl, Shubra Bansal); Gurugram Civil Hospital, Haryana, India (Umesh Mehta, Sunita Sharma, Brahmdeep Sindhu); Safdarjung Hospital, New Delhi, India (Sugandha Arya, Rekha Bharti, Harish Chellani, Pratima Mittal); Maulana Azad Medical College, New Delhi, India (Anju Garg, Siddharth Ramji), The Ultrasound Lab, Defence Colony, New Delhi, India (Ashok Khurana); Hamdard Institute of Medical Sciences and Research, Jamia Hamdard University, New Delhi, India (Reva Tripathi); All India Institute of Medical Sciences, New Delhi, India (Alpesh Goyal, Yashdeep Gupta, Smriti Hari, Nikhil Tandon); Government of Haryana, India (Rakesh Gupta); International Centre For Genetic Engineering and Biotechnology, New Delhi, India (Dinakar M Salunke Co-PI); G Balakrish Nair (Rajiv Gandhi Centre for Biotechnology, Trivandrum); Gagandeep Kang (Christian Medical College, Vellore).

## Additional files

Additional File 1: Supplementary information (PDF)

Additional File 2: Supplementary information tables (XLS)

