## Supplementary Information for "Comparison of first trimester dating methods for gestational age estimation and their implication on preterm birth classification in a North Indian cohort"

**SUPPLEMENTARY NOTE**

**Comparison of USG-Hadlock and LMP-based methods for estimation of GA in the first trimester**

To quantify the contribution of unreliable LMP, a sub-dataset was derived ($N_o$ = 1261 from $N_p$ = 791) from the TRAINING DATASET (Figure 1) by excluding participants with the use of contraceptives a month prior to the pregnancy, assisted conception, enrolment BMI beyond the normal range, and breastfeeding in the two months before conception.

Another analysis was done to estimate the contribution of foetal growth restriction to this discrepancy by creating a subset ($N_o$ = 1281 from $N_p$ = 820), filtered from the TRAINING DATASET by excluding participants with a known risk of foetal growth restriction. The factors that affect CRL measurements included active and passive smoking, consumption of tobacco and alcohol, and enrolment BMI beyond the normal range.

**Sensitivity analysis to test the effect of paired CRL measurements on performance of the dating model**

Since, for some participants, CRL was measured twice during their first trimester, we wanted to test the effect of these paired CRL measurements on the performance of Garbhini-GA1 formula. One can argue that more than one measurement of CRL of the same foetus in one sense is a dependent variable. However, we have considered them as separate observations to ensure our training data includes women whose CRL were measured in the latter part of the first trimester. Such improved coverages, we believe will improve the applicability of our formula even when a pregnant woman comes for her dating in the latter part of the first trimester. However, considering the dependence of paired measurements of CRL, we have now performed mixed modelling introducing a random effect term in the regression model. However, there was no difference between Garbhini-GA1 and the mixed-effect model [$GA\ (weeks)\ =\ 6.26769 + 1.53902(crl) - 0.07713(crl^2)$] as shown by BA analysis (see Table S12). Considering the simplicity of the formula, we have retained Garbhini-GA1 as our primary first-trimester GA dating formula.

**SUPPLEMENTARY FIGURES**

**Figure S1: The simulated data points using the Hadlock equation (blue) added to points from the (A) TRAINING, (B) DBSCAN and (C) CLINICALLY-FILTERED DATASET shown in red.** The x-axis is CRL in cm, and the y-axis is LMP-based GA in weeks.

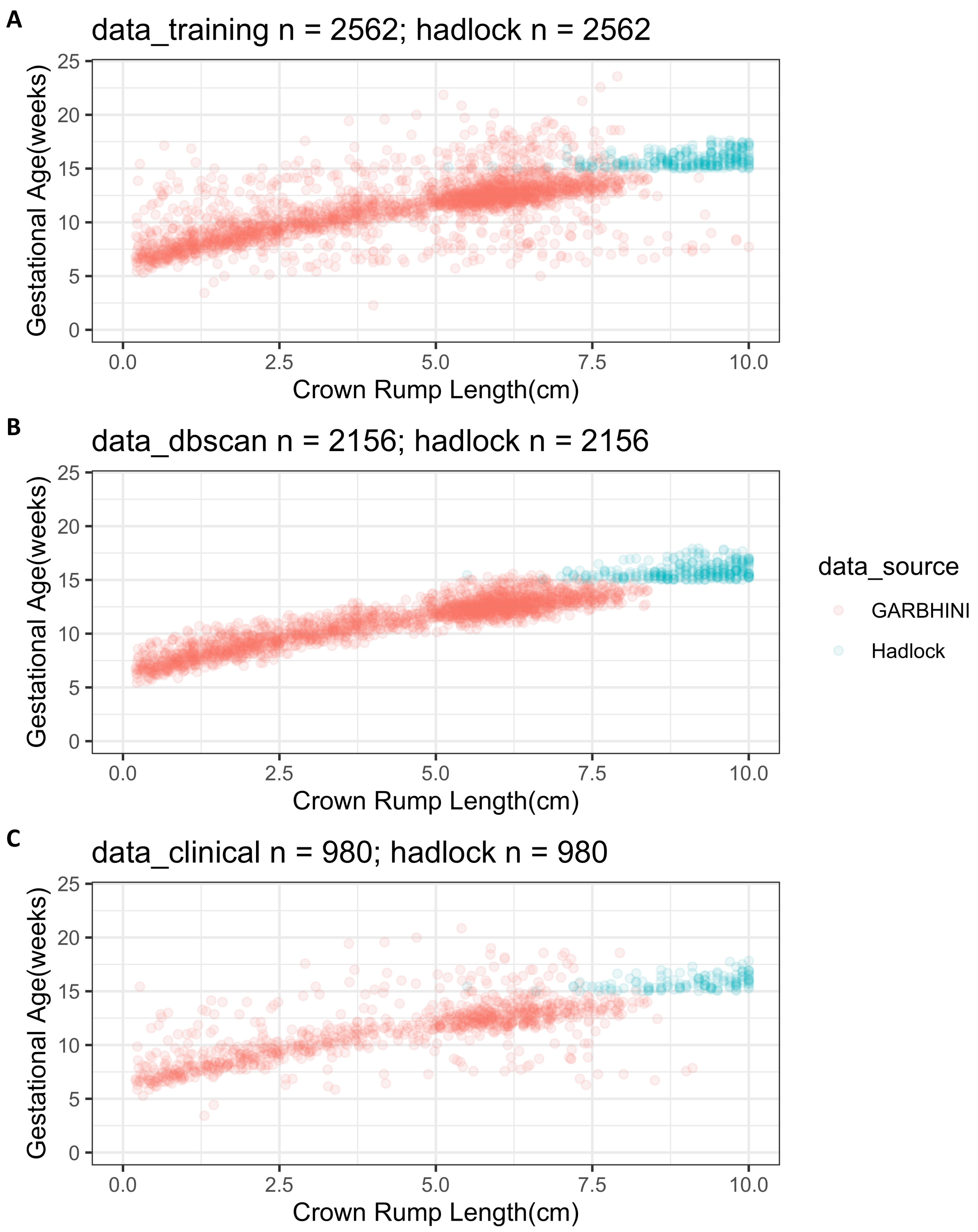

**Figure S2: Top performing fractional regression lines on the (A) TRAINING, (B) DBSCAN and (C) CLINICALLY-FILTERED DATASET with the Hadlock data supplementation on each.** The x-axis is CRL in cm, and the y-axis is LMP-based GA in weeks. Regression lines are shown for the different polynomial fits, as are the points selected after filtering.

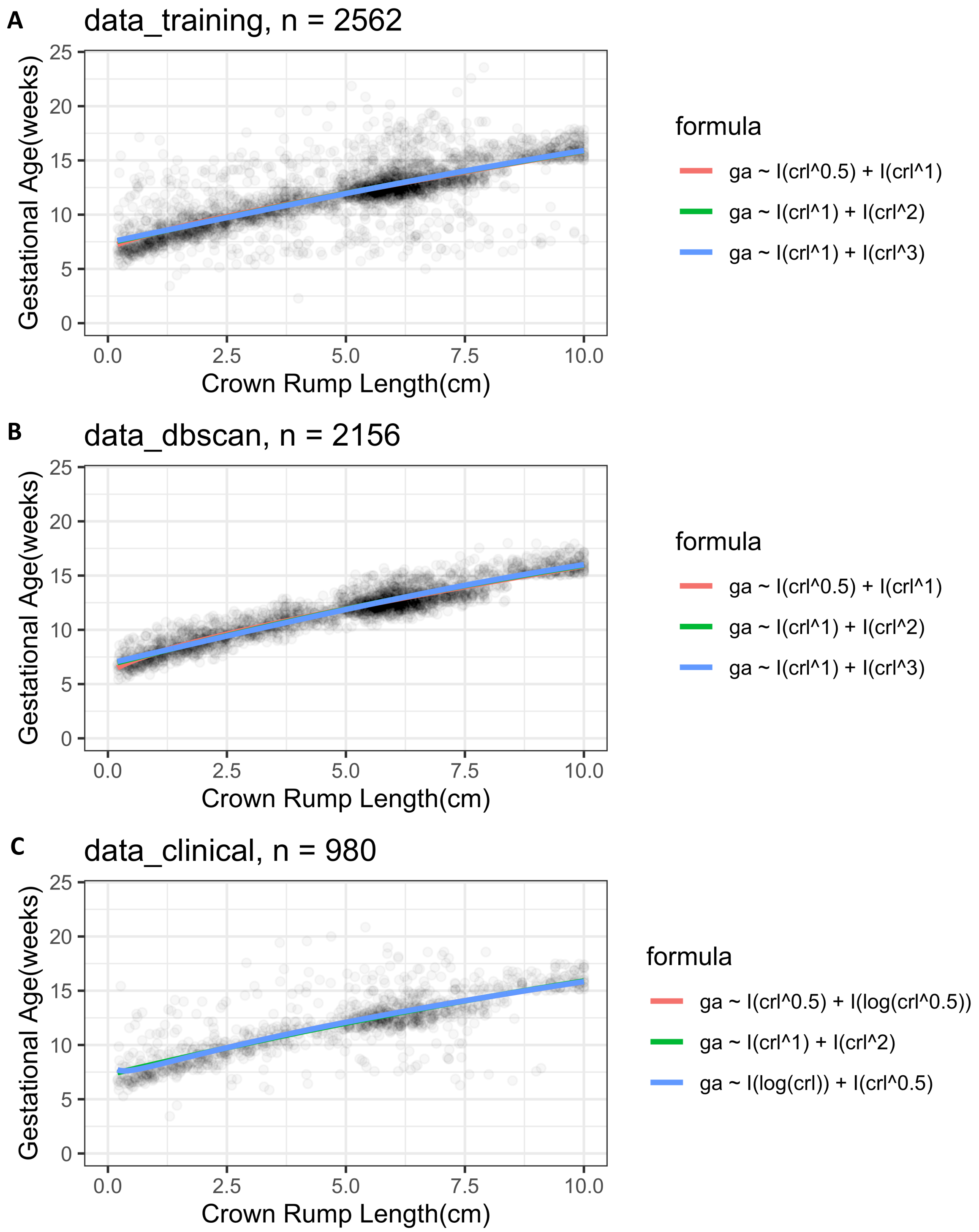

**Figure S3: Distribution of errors for generated multivariable formulae.** The x-axis is the error (the difference between predicted and LMP-based GA), the y-axis is the density for each subplot, with formulae. The variable description used in the formulae is given Table S4.

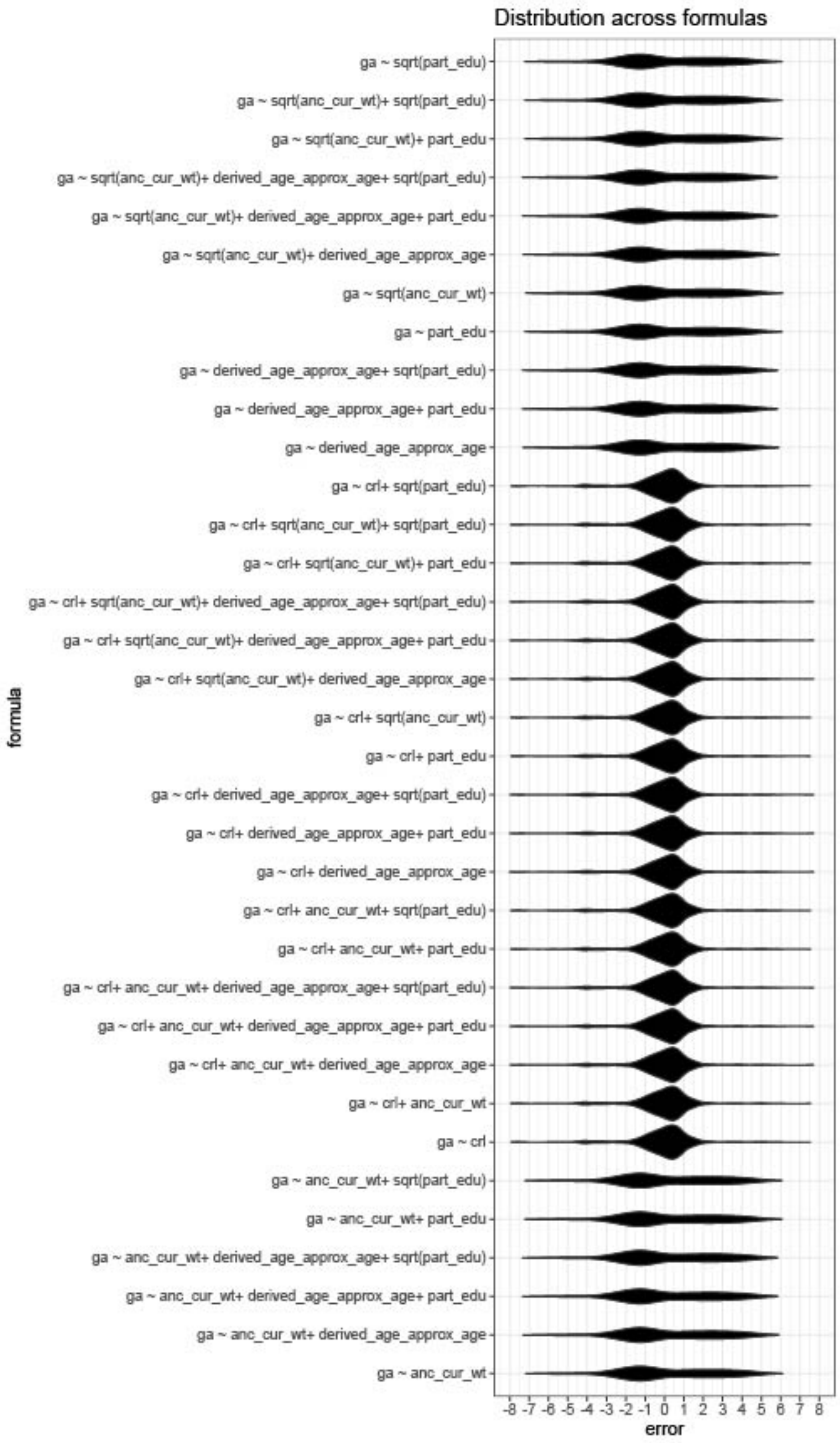

**Figure S4: (A)** Fit of published formulae, Garbhini-GA1 on the TEST dataset; x-axis is CRL in cm, the y-axis is GA in weeks (predicted are lines, LMP-based are datapoints); **(B)** Distribution of errors for published formulae, Garbhini-1 on TEST dataset; x-axis is the error (the difference between predicted and LMP-based GA), the y-axis is the density for each subplot, with formulae names.

A

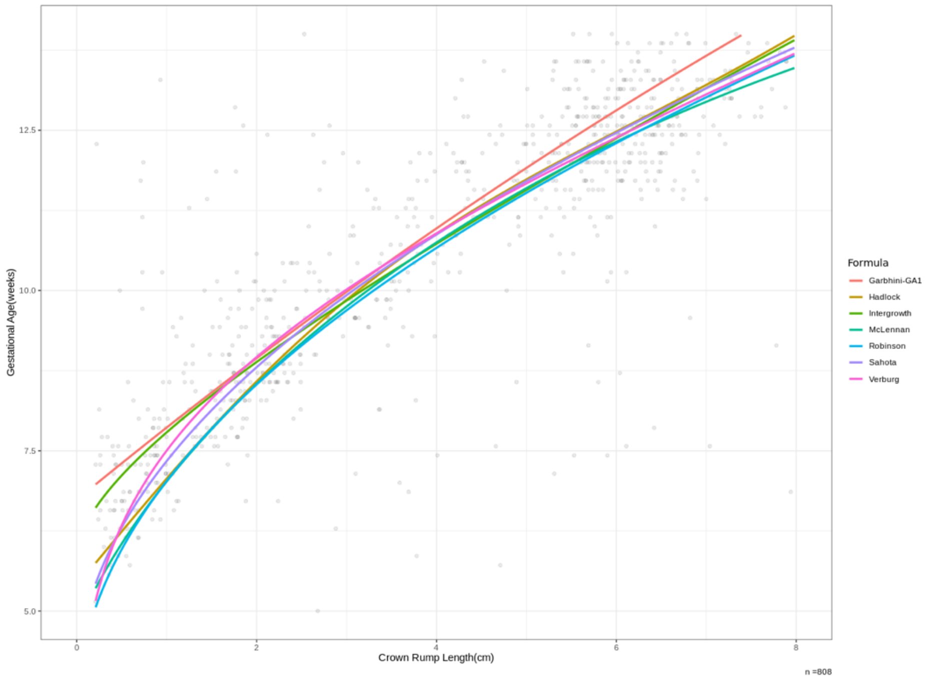


B

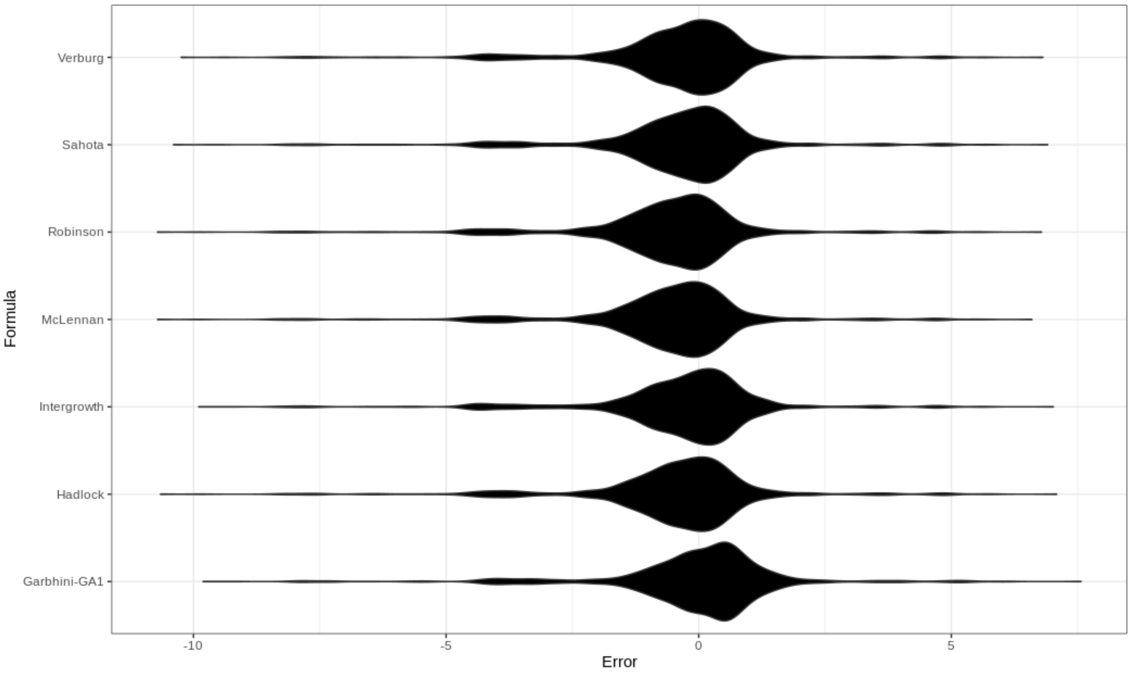

**Figure S5:** Examples of USG-based determination of CRL.

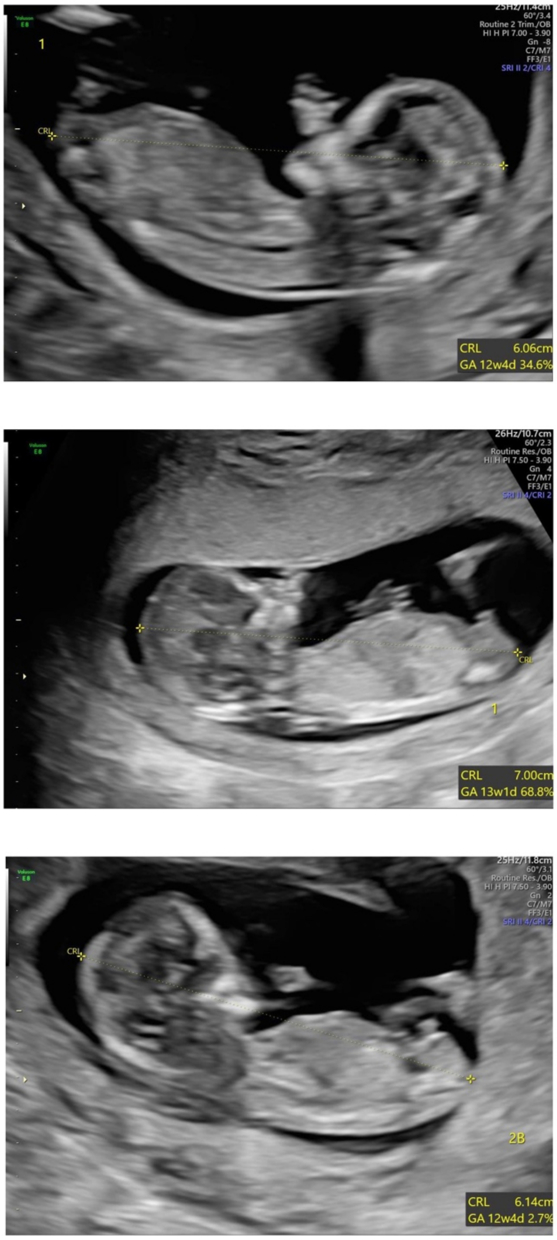

**SUPPLEMENTARY TABLE LEGENDS**

**Table S1:** The 'high quality' published dating formulae evaluated in this study along with the population and number of participants used to develop the formula. GA is estimated either in days or weeks as indicated. Crown-rump length is measured in cm in all formulae, indicated as *CRL*. For INTERGROWTH–21$^{st}$, the crown-rump length is measured in mm and is indicated as *crl*.

**Table S2:** Selection criteria for clinical filtering to identify participants serving as a reference for dating

**Table S3:** Number of samples selected, and the number of clusters formed when applying DBSCAN to remove noise. The columns represent the distance threshold for counting neighbours, and the rows represent the number of neighbours required to be classified as a cluster core-point. The cells are n, c where n is the number of data points after filtering, and c is the number of clusters formed

**Table S4:** Features related to gestation in the first trimester potentially predictive of GA

**Table S5:** Features selected using linear modelling and random forest selection

**Table S6:** Adjusted $R^2$, RMSE values multivariable models with an additional column for adjusted $R^2$ on the TRAINING dataset.

**Table S7:** $R^2$, RMSE values fractional polynomial models on the TRAINING dataset supplemented with datapoints generated using the Hadlock formula.

**Table S8:** Adjusted $R^2$, RMSE values for published formulae, and Garbhini-GA1 on the TEST dataset

**Table S9**: Pairwise comparison of preterm births classified by different formulae using Fisher's Exact test followed by Bonferroni p-value correction.

**Table S10:** Indicators of performance of different formulae in the classification of preterm birth.

**Table S11:** Counts for numbers of participants in TRAINING dataset with the given gestational age in weeks.

**Table S12:** Pairwise comparison of mean difference (LoA) between different first-trimester dating formulae (Difference: Column formula - Row formula) for the TEST DATASET ($N_o$ = 808).
